# Large Language Model Embeddings of Surgical Procedural Names for Confounding Adjustment in Perioperative Observational Studies

**DOI:** 10.64898/2026.08.22.26361065

**Authors:** Lichy Han, Marc Ghanem, Mahitha K. Simhambhatla, Philip Chung, Nima Aghaeepour

**Affiliations:** Department of Anesthesiology, Perioperative and Pain Medicine, Stanford University School of Medicine, Stanford, CA; The University of Texas at Austin, Austin, TX; Department of Pediatrics, Stanford University School of Medicine, Stanford, CA; Department of Biomedical Data Science, Stanford University School of Medicine, Stanford, CA

## Abstract

**Background:** Perioperative observational studies are increasingly used to evaluate anesthesia practices that are difficult to test in randomized trials, but treatment selection is influenced by surgical procedure type. Scalable and robust methods are needed to adjust for procedure-level confounding across heterogeneous surgical cohorts.

**Methods:** We developed and assessed the utility of a surgical-name embedding framework using free-text procedure names from 627,624 adult perioperative records. Procedure names were embedded using open-source sentence-embedding models, reduced with principal components analysis, and incorporated into entropy-balanced observational analyses. We compared unweighted, clinical covariate adjustment, and clinical covariate plus surgical-name embedding adjustment across three replications of recent perioperative randomized trials: GA-CARES for total intravenous versus volatile anesthesia on cancer mortality, PADDI for dexamethasone and surgical site infection, and GAP for perioperative gabapentin and postoperative length of stay.

**Results:** In the GA-CARES replication, unweighted and clinical covariate adjustment suggested lower two-year mortality with total intravenous anesthesia, whereas adding surgical-name embeddings attenuated the estimate to a nonsignificant association consistent with the randomized trial (OR 0.84 [95% CI, 0.62-1.14]; p=0.274). In the PADDI replication, adding surgical-name adjustment reproduced the trial’s overall non-harm finding for 30-day surgical site infection (OR 0.81 [95% CI, 0.77-0.85], p<0.001) while preserving the expected protective association with postoperative nausea and/or vomiting. In the GAP replication, the full cohort was null across adjustment strategies, but surgical-name embeddings were required to recover trial-consistent null length-of-stay estimates across cardiac, thoracic, and abdominal subgroups. Department indicators and randomly generated covariates did not reproduce the effect of surgical-name embeddings, supporting the presence and importance of procedure-specific information.

**Conclusions:** Free-text surgical-name embeddings provide a scalable method for representing surgical context in perioperative observational studies. Surgical-name adjustment improved concordance with randomized trial benchmarks while preserving expected treatment effects, supporting its use as an additional layer of confounding adjustment in large perioperative datasets.

## Introduction

With over 300 million surgical procedures that are performed annually worldwide,^1^ the increasing volume in parallel with the digitization of perioperative records has created large observational datasets capable of studying anesthesia decisions that are difficult to test in randomized trials.^2^ However, treatment selection in anesthesia is closely tied to the surgical procedure. Surgical procedure type may influence anesthetic technique, adjunctive medication use, baseline outcome risk, and postoperative recovery. Therefore, inadequate adjustment for procedure-level differences can bias observational estimates.^3^

To address indication bias, perioperative observational studies often restrict analyses to a small set of procedures,^4,5^ perform multiple subgroup analyses,^6^ or exclude large groups of cases altogether.^7^ These strategies can reduce confounding but may also limit sample size, generalizability, and statistical power. Studies that do adjust for procedure typically use procedural codes, service lines, or manually defined procedure groups.^7–9^ These approaches may be too coarse for heterogeneous perioperative cohorts and may not capture complex operations with multiple procedural components. Procedures within the same specialty can differ substantially in expected duration, contamination risk, pain trajectory, blood loss, and length of stay, whereas procedures performed by different specialties may share clinically relevant features.^10,11^ These limitations create a need for scalable methods that represent procedure context more flexibly than standard structured categories.

Free-text operative procedure names are routinely available in perioperative records and contain information about surgical site, approach, laterality, revision status, and procedural complexity. Large language model embeddings provide a way to convert procedure names into continuous features that preserve semantic similarity and can be incorporated into causal models as procedure-level covariates.^12^

We evaluated whether surgical-name embeddings improve confounding adjustment in perioperative observational analyses. We assessed multiple sentence-embedding large language models (LLMs) with dimensionality reduction and added the resulting surgical-text features to entropy-balanced^13^ analyses across three observational replications of recent randomized trials. These trial benchmarks included propofol total intravenous anesthesia versus volatile anesthesia in cancer surgery (GA-CARES),^14^ dexamethasone and surgical site infection (PADDI),^15^ and perioperative gabapentin and postoperative length of stay (GAP).^16^ We hypothesized that incorporating surgical-name embeddings would improve adjustment for procedure-level confounding and produce observational estimates more concordant with randomized trial results.

**Figure 1.**
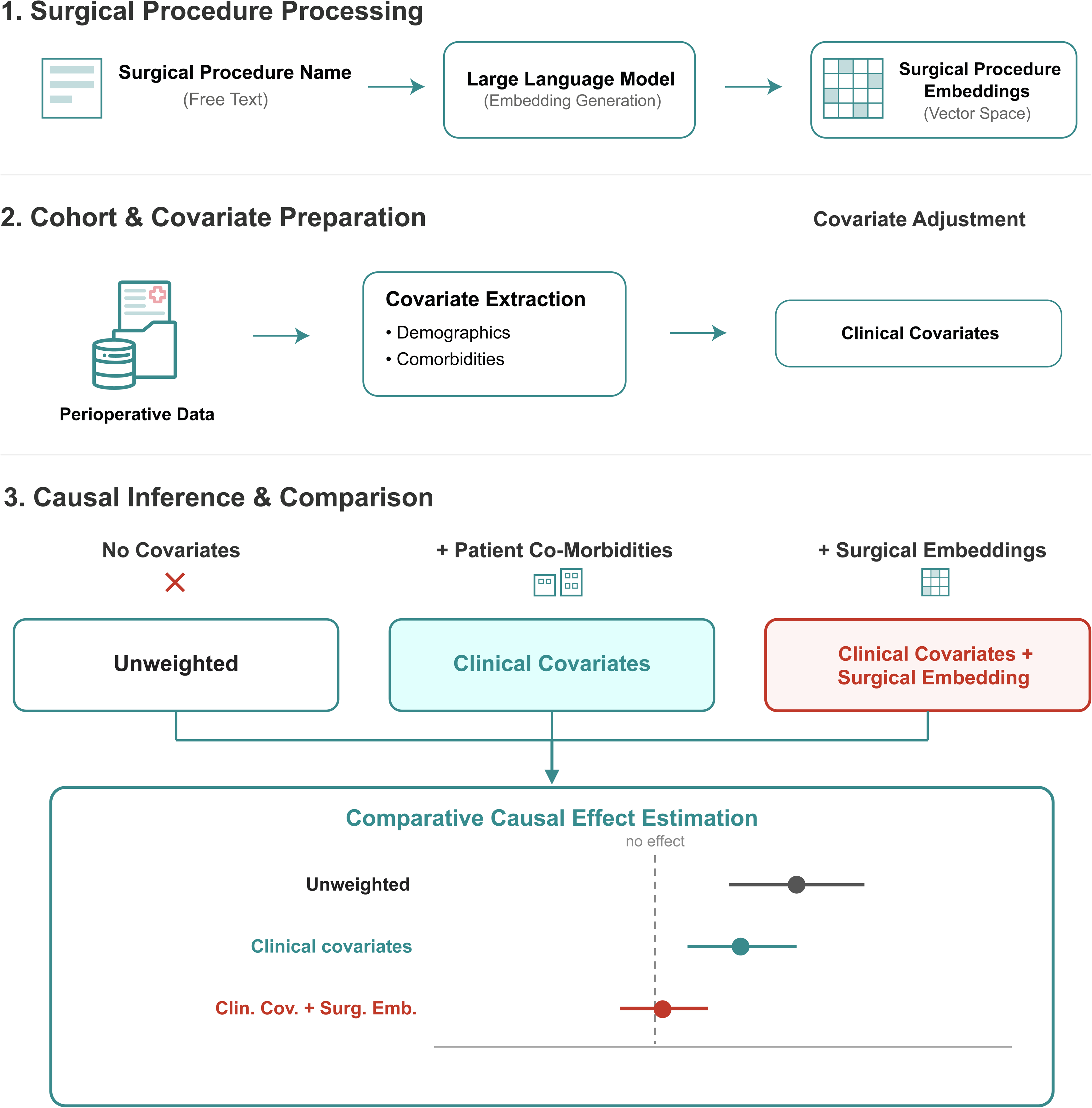
LLM surgical embedding framework for confounding adjusted causal inference in perioperative observational studies. Free-text surgical procedure names are processed through a three-stage analytical pipeline that compares confounding adjustment strategies across clinical applications. **(1)** Surgical Procedure Processing: free-text operative procedure names are passed through a pre-trained biomedical large language model (LLM) to generate dense vector embeddings of surgical context, which are included as continuous covariates in adjusted models. **(2)** Cohort and Covariate Preparation: perioperative patient-level data are extracted for each application, and patient demographics and comorbidities are assembled into a clinical covariate set for adjustment. **(3)** Causal Inference and Comparison: treatment-effect estimates are compared across three adjustment strategies: unweighted (no covariates), clinical covariates (demographics plus comorbidities), and clinical covariates plus surgical-name embeddings. The trajectory of estimated effects (illustrative forest plot) shows the degree to which surgical-name embeddings attenuate residual confounding beyond standard covariate adjustment.

## Materials and Methods

### Study Design

We conducted a retrospective, trial-benchmarked observational study to evaluate whether free-text surgical-name embeddings derived using large language models improve confounding adjustment in perioperative causal analyses. This study was approved by the Stanford University Institutional Review Board (protocol 88311). A data analysis and statistical plan was written after the data were accessed.

We first identified free-text operative procedure names from the perioperative record and used pre-trained sentence-embedding models to convert each procedure name into a dense numerical representation of surgical context. These embeddings can capture clinical information, including anatomic site, operative complexity, and procedure-specific treatment patterns that may not be represented by structured groups or codes. In parallel, patient-level perioperative data were assembled for each randomized-trial replication, including demographics, clinical comorbidities, treatment exposures, and study-specific outcomes. We then compared treatment-effect estimates across three analytic strategies: unweighted analyses, entropy balancing using structured clinical covariates, and entropy balancing using structured clinical covariates plus surgical-name embedding components. Entropy balancing was selected because it directly imposes covariate balance constraints, avoiding reliance on correctly specified propensity-score models.^13^ Results from all three strategies were compared with the corresponding randomized trial benchmarks to assess whether surgical-name embeddings improved concordance with the trial results.

### Perioperative Data Source

Our cohort consisted of 627,624 adult surgical cases from Stanford University from November 10, 2012 to April 25, 2026. Each clinical replication applied study-specific eligibility criteria, exposure definitions, outcomes, and follow-up windows aligned as closely as possible to the corresponding randomized trial. Cohort flow and eligibility criteria for the three replications are shown in Figure 2.

**Figure 2.**
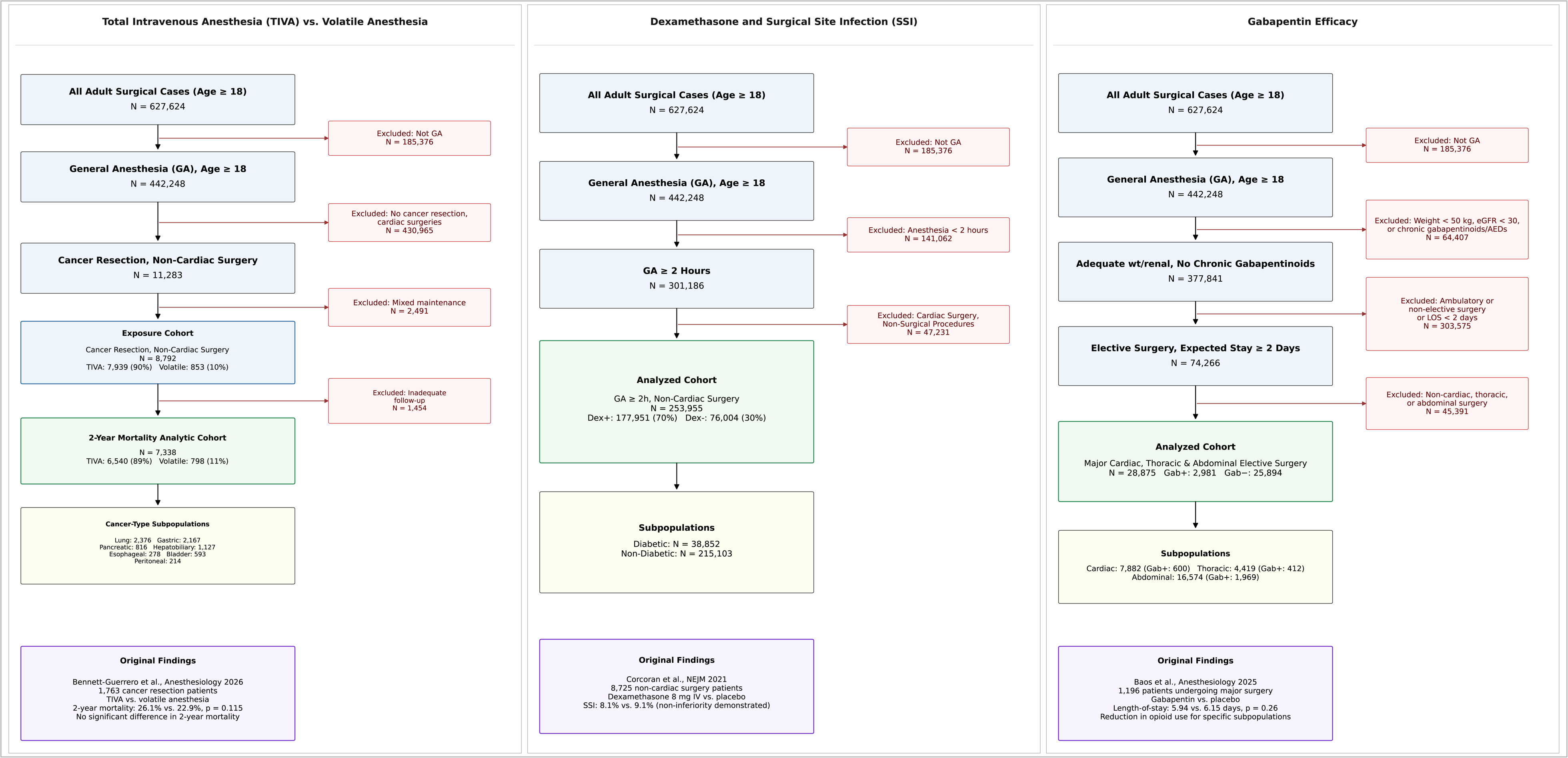
Combined cohort flow diagrams for the three perioperative observational study cohorts replicated in this study, each derived from a common source population of 627,624 adult surgical cases. Left column: total intravenous anesthesia (TIVA) versus volatile anesthesia and cancer mortality. After restriction to general anesthesia, non-cardiac cancer-resection surgery, and exclusion of mixed-maintenance cases, the analyzed cohort comprised 7,338 patients (TIVA 6,540 [89%]; volatile 798 [11%]), primary outcome is 2-year mortality, replicating the GA-CARES trial (Bennett-Guerrero et al., Anesthesiology 2026). Center column: dexamethasone and surgical site infection (SSI). After restriction to general anesthesia lasting ≥2 hours and exclusion of cardiac and non-surgical procedures, the analyzed cohort comprised 253,955 non-cardiac cases (dexamethasone-treated 177,951 [70%]; untreated 76,004 [30%]), with diabetic (N=38,852) and non-diabetic (N=215,103) subpopulations; primary outcome is 30-day SSI, replicating the PADDI trial (Corcoran et al., NEJM 2021). Right column: gabapentin efficacy. After exclusion of patients with low weight, renal impairment, or chronic gabapentinoid use and restriction to elective surgery with expected stay ≥2 days in cardiac, thoracic, or abdominal specialties, the analyzed cohort comprised 28,875 patients (gabapentin-exposed 2,981; unexposed 25,894); primary outcome is hospital length of stay, replicating Baos et al. (Anesthesiology 2025). Exclusion criteria (red), analyzed cohorts (green), subpopulations (yellow), and original trial findings (purple) are shown for each analysis.

### Surgical-Name Embedding Pipeline

For each surgical procedure, the free-text operative procedure name was passed through a pre-trained, open-source large language sentence-embedding model to generate dense vector representations. We tested three different models: BGE-large (1024 dimensions), MiniLM (384 dimensions), and SapBERT (768 dimensions). BGE-large is a general-purpose embedding model from the Beijing Academy of Artificial Intelligence (BAAI) General Embedding (BGE) family,^17^ MiniLM is a compact distilled transformer sentence model,^18^ and SapBERT is a biomedical entity-representation model trained with self-alignment pretraining (Sap)^19^ based on the original Bidirectional Encoder Representations from Transformers (BERT) models.^20^ These three models were selected to allow us to evaluate whether surgical procedure names were better represented by broad semantic similarity or domain-specialized biomedical language structure. Because raw embedding vectors are high-dimensional and often contain correlated features, we used principal components analysis (PCA) to reduce dimensionality before causal modeling, retaining a lower-dimensional representation that preserved dominant variation in surgical procedure semantics while improving model stability and covariate balance.

### Clinical Covariate Selection

Clinical covariates were selected *a priori* based on the covariates used by the National Surgical Quality Improvement Program (NSQIP), a widely used surgical outcomes registry and risk-adjustment framework.^8^ Across replications, these included demographics, body mass index, American Society of Anesthesiologists physical status,^21,22^ and major comorbidities including diabetes, hypertension, congestive heart failure, chronic obstructive pulmonary disease, renal dysfunction, ascites, and disseminated cancer. We then added the MiniLM PCA-68 surgical-name embedding components to these structured clinical covariates to assess whether free-text surgical procedure context improved balance beyond conventional clinical risk adjustment.

### Randomized-Controlled Trial Replications: Total Intravenous Anesthesia vs. Volatile Anesthesia

The cancer-surgery analysis was designed to mirror the General Anesthetics in Cancer Resection study (GA-CARES), a multicenter, partially blinded randomized superiority trial motivated by the hypothesis that propofol-based anesthesia might improve cancer outcomes by preserving immune-mediated clearance of circulating tumor cells.^14^ The original trial enrolled adults undergoing high-risk oncologic resections and randomized patients 1:1 to propofol or volatile anesthetic maintenance for general anesthesia. Our observational replication compared propofol total intravenous anesthesia (TIVA) with volatile anesthetic maintenance among cancer-resection cases identified by procedure-name regular expressions. TIVA was defined as general anesthesia with propofol maintenance and without volatile exposure; volatile anesthesia was defined as general anesthesia with volatile maintenance. Mixed propofol and volatile cases were excluded. Only non-cardiac surgeries were included, and subgroups included lung, gastric, pancreatic, hepatobiliary, esophageal, bladder, and peritoneal procedures. The trial’s primary outcome was 2-year mortality.

### Randomized-Controlled Trial Replications: Dexamethasone and Surgical Site Infection

The infection analysis was designed to mirror the original Perioperative Administration of Dexamethasone and Infection (PADDI) study, a randomized, placebo-controlled, triple-blind noninferiority trial testing whether a single antiemetic dose of dexamethasone increased surgical site infection (SSI).^15^ The original trial enrolled adult patients undergoing elective, non-urgent non-cardiac surgery under general anesthesia with expected operative duration of at least two hours and expected postoperative stay of at least one night. Patients were randomized 1:1, stratified by center and diabetes status, to 8 mg intravenous dexamethasone or placebo after induction and before incision. The primary outcome was surgical site infection within 30 days, with superficial, deep, and organ-space infections analyzed as secondary outcomes and postoperative nausea and/or vomiting (PONV) expected to improve with dexamethasone. Our observational replication used a PADDI-style cohort of general anesthetic, non-cardiac operations lasting at least 120 minutes, and accounted for antibiotic exposures. Outcomes included 30-day SSI, 90-day SSI, superficial, deep, and organ-space SSI, and PONV, defined as greater than 1 rescue antiemetic administration in the post-anesthesia care unit.

### Randomized-Controlled Trial Replications: Gabapentin Efficacy

The third study replicated was the Gabapentin for Pain Management after Major Surgery (GAP) study, a placebo-controlled, randomized trial testing gabapentin as an adjunct with major surgery.^16^ The original trial enrolled adults undergoing non-emergent cardiac, thoracic, or abdominal surgery who were expected to remain hospitalized for at least two postoperative days. Patients with antiepileptic or gabapentinoid use, estimated glomerular filtration rate less than 30 ml/min/1.73 m^2^, or weight less than 50 kg were excluded. The trial primary outcome was length of hospital stay, and secondary outcomes included pain scores and adverse events. Our observational replication used a GAP-style perioperative gabapentin cohort among adult general anesthetic cases with weight at least 50 kg, estimated glomerular filtration rate at least 30 ml/min/1.73 m^2^ when available, no pregabalin or antiepileptic exposure, non-emergent surgery, and postoperative length of stay at least two days. Treatment was preoperative gabapentin. Subgroups included cardiac, thoracic, and abdominal major surgery, mirroring the original study. Outcomes included length of stay and postoperative pain.

### Feature Comparison Analysis with Surgical Department and Random Controls

To evaluate whether the benefit of surgical-name embeddings reflected meaningful surgical-text information rather than broad service labels or nonspecific high-dimensional adjustment, we performed a feature-comparison analysis. For each trial replication, treatment-effect estimates were compared across clinical-covariate adjustment alone and clinical covariates augmented with one of four surgical-context feature sets: structured surgical department indicators using 1-hot encoding, randomly generated covariates matched to the dimensionality of the embedding representation, randomly scrambled surgical-name embedding components, or the actual surgical-name embedding components. Surgical department indicators tested whether broad service-line information was sufficient to account for surgical context, randomly generated covariates tested whether adding model dimensions alone could reproduce the embedding effect, and scrambled embeddings tested whether the observed results required preservation of the relationship between surgical names and their embedding values.

### LLM and Dimensionality Reduction Sensitivity Analysis

To assess the dependence of the surgical-text adjustment on LLM choice and the amount of embedding variance retained, we conducted two sensitivity analyses on representation specification. First, we repeated the surgical-name embedding adjusted workflow across the three sentence-embedding models, all at 75% variance. Second, we tested the sensitivity to the number of dimensions retained using MiniLM by varying the number of principal components used from the surgical-name embeddings. Each of these analyses was performed for all three studies for all primary outcomes.

### Statistical Analysis

All analyses were performed using Python 3.11.15. Embeddings were generated with sentence-transformers 6.0.0 and PyTorch 2.12.0. For each application, entropy balancing was used to balance either clinical covariates alone or clinical covariates plus surgical-name embedding components. Binary outcomes were analyzed using weighted logistic regression and used odds ratios with 95% confidence intervals. Continuous outcomes were analyzed using weighted log-linear models and used geometric mean ratios. We compared unweighted, clinical covariate adjusted, and clinical covariate plus surgical-name adjusted estimates against randomized trial results. Embeddings were visualized using uniform manifold approximation and projection (UMAP).^23^ Statistical significance was defined as p<0.05.

## Results

### Cohort Characteristics

We derived three cohorts from our perioperative data to match the three randomized controlled trials (RCTs), and baseline characteristics, exposure distributions, and outcomes are detailed in Tables S1-S3. In the GA-CARES replication assessing propofol TIVA versus volatile anesthesia, patients differed in demographic profile, cancer procedure mix, and baseline survival risk, consistent with nonrandom anesthetic selection across oncologic surgical programs (Table S1). In the PADDI cohort assessing dexamethasone and surgical site infection, dexamethasone-treated patients differed from untreated patients in comorbidity burden, diabetes-related risk, and crude infection and antiemetic outcome rates, suggesting both patient-level and procedure-level channeling of steroid use (Table S2). Antibiotic exposures were also taken into account for the PADDI analysis (Table S4). In the GAP replication cohort studying gabapentin, patients given gabapentin differed from unexposed patients by body mass index, renal function, comorbidities, surgical subgroup, and postoperative pain profiles (Table S3). Across all three analyses, exposure groups differed in patient characteristics, comorbidity burden, procedural composition, and outcome prevalence, motivating covariate balancing before treatment-effect estimation.

### LLM Embeddings of Surgical Procedure Names

The surgical-name embedding analyses supported MiniLM with PCA-based dimensionality reduction as the primary representation for downstream modeling. All three LLM models tested (BGE-large, MiniLM, SapBERT) produced broadly similar surgical embedding structure when visualized by surgical department (Figure S1A-C). When assessing dimensionality reduction with PCA, SapBERT required the least number of dimensions to retain 75% variance but had the lowest cluster quality (Figure S1D-F), whereas BGE-large and MiniLM were comparable in variance retention and cluster quality (Figure S1E, S1F). Because MiniLM provided similar structure with a lower-dimensional raw embedding space, it was selected for downstream analysis. In the cumulative variance plot, 75% retained variance corresponded to the elbow of the curve and was selected as the primary dimensionality-reduction target. We then assessed MiniLM specifically across varying degrees of dimensionality reduction (Figure S2A-F). At 75% retained variance, the overall embedding structure remained well preserved, with distinct procedural clusters (Figure S2C). Closer inspection showed that surgical departments often formed distinct regions, while related procedures also clustered across departmental boundaries, including abdominal and pelvic operations spanning multiple departments and spine procedures from both orthopedic surgery and neurosurgery (Figure S3). Together, these analyses supported the use of PCA-reduced MiniLM embeddings as a computationally efficient, clinically coherent representation.

### GA-CARES: Anesthetic Maintenance and Cancer Mortality

The GA-CARES replication recovered the randomized trial’s null overall survival finding only with the use of surgical-name embeddings for covariate adjustment. The original GA-CARES trial found no survival advantage for propofol TIVA compared with volatile anesthetic maintenance at two years. In our observational cohort, propofol TIVA and volatile anesthesia were distributed unevenly across cancer types (Figure 3A), and crude two-year mortality differed by cancer subgroup (Figure 3B). In treatment-effect analyses, both the unweighted analysis and adjustment with clinical covariates alone suggested significantly lower two-year mortality with TIVA, contradicting the randomized trial benchmark (Figure 3C). After adding MiniLM PCA-68 surgical-name embeddings, the all-cancer estimate attenuated to a nonsignificant association (OR 0.84 [95% CI, 0.62-1.14]; p=0.274), aligning with the randomized trial result (Figure 3C). The corresponding survival curves show the same pattern across adjustment strategies, with an apparent TIVA survival advantage in the unweighted and clinical covariate adjusted analyses that was attenuated after surgical-name adjustment (Figure 3D-F). Together, these findings suggest that surgical procedure context was an important source of confounding in the observational GA-CARES replication and was necessary to recover the trial-consistent null overall survival result.

**Figure 3.**
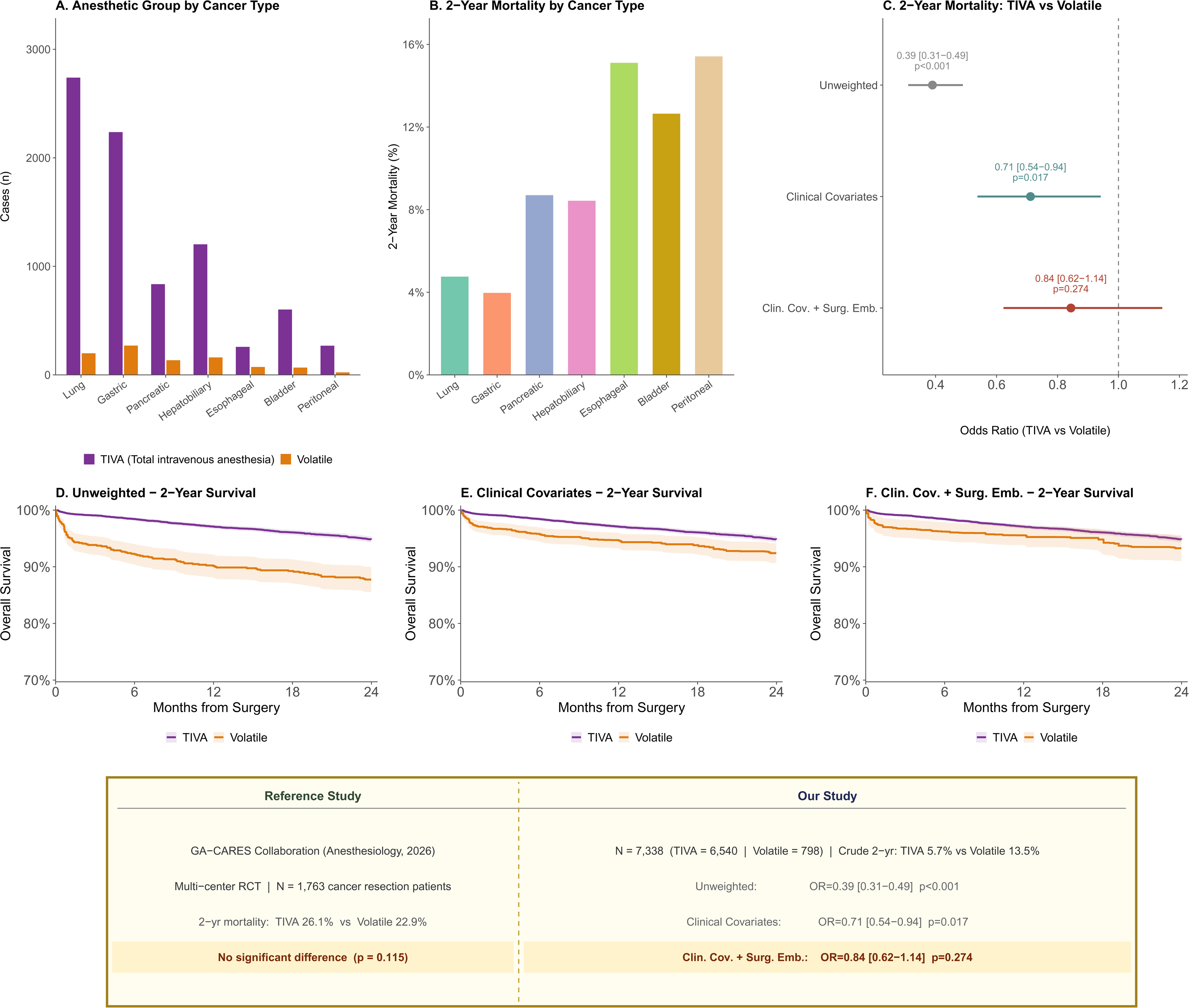
LLM surgical embeddings attenuate a spurious protective association between propofol total intravenous anesthesia and cancer mortality, consistent with the null result of the GA-CARES randomized trial. The GA-CARES trial (Bennett-Guerrero et al., Anesthesiology 2026) randomized 1,763 patients undergoing cancer resection to propofol TIVA versus volatile inhalational maintenance and found no survival difference at 2 years (26.1% vs. 22.9% mortality; p=0.115). **(A)** Case volume by cancer type, grouped by anesthetic type (TIVA, purple; volatile, orange). Cancer-type categories were non-mutually exclusive. **(B)** 2-year mortality by cancer type, ranging from ∼4% (gastric) to ∼15% (esophageal, peritoneal). **(C)** Forest plot of odds ratios (TIVA vs. volatile) for 2-year mortality across three strategies: unweighted (0.39 [0.31– 0.49], p<0.001), clinical covariates (0.71 [0.54–0.94], p=0.017), and clinical covariates plus surgical-name embeddings (0.84 [0.62–1.14], p=0.274); the estimate moves toward the null and loses significance as adjustment increases. **(D–F)** Covariate-balanced weighted Kaplan–Meier 2-year overall survival curves (TIVA, purple; volatile, orange) under unweighted (D), all clinical covariates (E), and all clinical covariates plus surgical-name embeddings (F) adjustment; the apparent TIVA survival advantage is progressively attenuated after clinical covariate and surgical-name embedding adjustment. The summary panel contrasts the reference trial with our replication across the three strategies.

### PADDI: Dexamethasone and Surgical Site Infection

In the dexamethasone replication, surgical-name adjustment preserved PADDI’s expected noninferiority, and did not indiscriminately attenuate all associations to null. SSI rates by wound depth and time point are shown in Figure 4A, with diabetes prevalence by SSI category shown in Figure 4B, and the difference in dexamethasone usage on PONV in Figure 4C. As in PADDI, the primary any-SSI analysis did not show evidence of increased infection risk with dexamethasone; in the MiniLM PCA-68 embedding adjusted analysis, dexamethasone was associated with lower odds of any SSI at both 30 days (OR 0.81 [95% CI, 0.77-0.85], p < 0.001) and 90 days (OR 0.81 [95% CI, 0.78-0.85], p < 0.001; Figure 4D). When analyzing SSI subtypes, deep and organ-space SSI were null, consistent with the original trial’s finding of similar deep and organ-space infection rates, whereas superficial SSI was higher with dexamethasone (OR 1.32 [95% CI, 1.14-1.52], p < 0.001; Figure 4D). The change in the 30-day any-SSI estimate across adjustment strategies is shown in Figure 4E.

**Figure 4.**
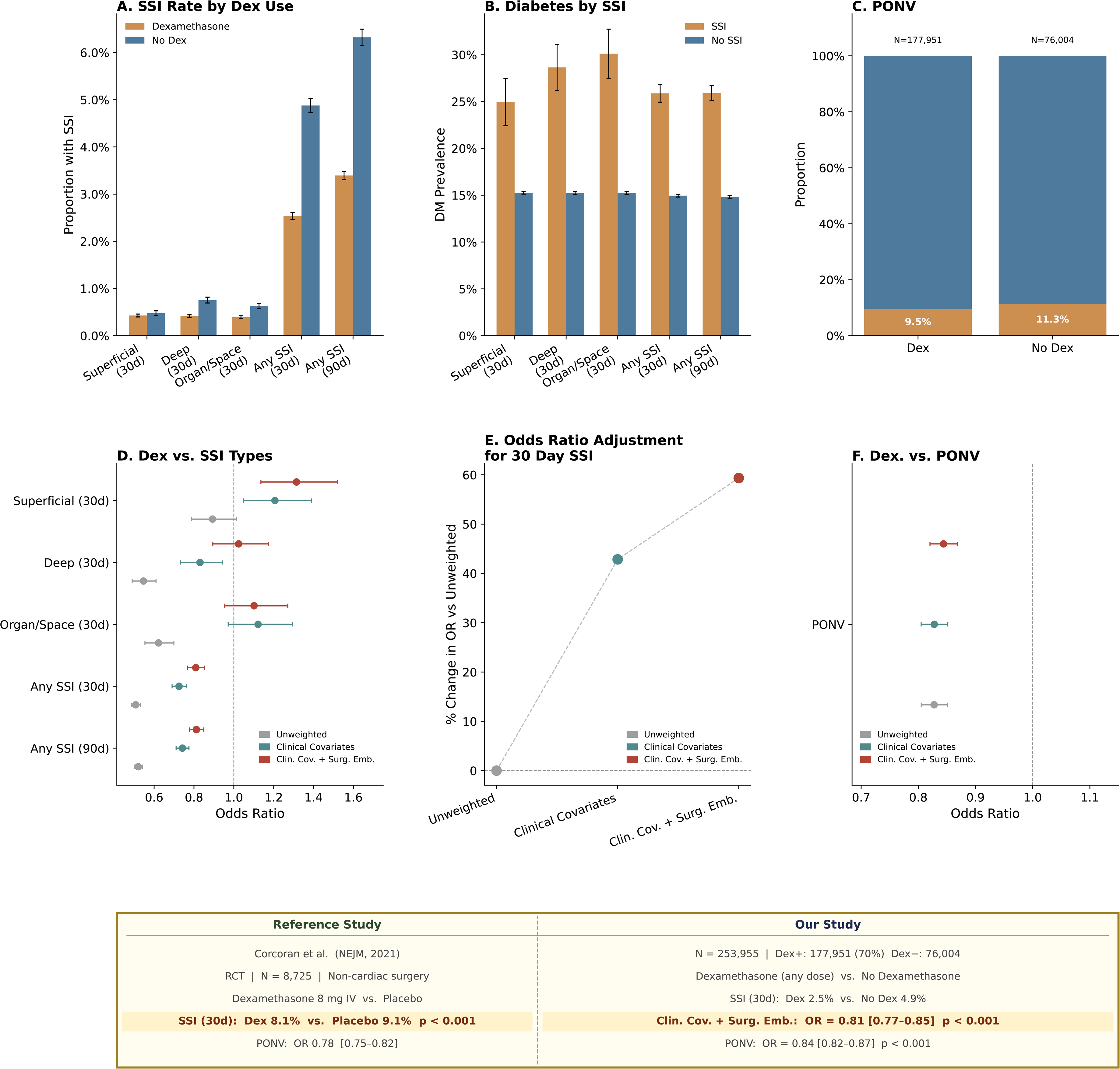
LLM surgical embeddings maintain the non-harmful association between dexamethasone and surgical site infection, consistent with the noninferiority finding demonstrated by the PADDI randomized controlled trial. The PADDI trial (Corcoran et al., NEJM 2021) randomized 8,725 patients undergoing non-cardiac surgery to intraoperative dexamethasone 8 mg IV versus placebo and demonstrated non-inferiority for SSI (8.1% vs. 9.1%). **(A)** Raw SSI rates by wound depth and time point (superficial, deep, organ/space at 30 days; any SSI at 30 and 90 days), stratified by dexamethasone use (treated, tan; untreated, blue). **(B)** Diabetes mellitus prevalence stratified by SSI occurrence across the same categories (SSI, tan; no SSI, blue), confirming enrichment of metabolic comorbidity among infected patients. **(C)** Postoperative nausea and/or vomiting (PONV) by dexamethasone use (9.5% vs. 11.3%), an internal positive control for dexamethasone’s antiemetic effect. **(D)** Forest plot of odds ratios for dexamethasone versus SSI by wound depth and time point across unweighted, clinical covariate adjustment, and clinical covariate plus surgical-name embedding adjustment. Dexamethasone continued to be associated with lower odds of overall SSI at both 30 and 90 days after full adjustment. Depth-specific estimates were heterogeneous, with no significant association for deep or organ-space SSI after surgical-name adjustment and higher odds of superficial SSI. This pattern is broadly consistent with the PADDI trial’s overall noninferiority finding while suggesting a subtype-specific superficial SSI signal in this observational cohort. **(E)** Percent change in the 30-day any-SSI odds ratio relative to the unweighted estimate, rising from 0% (unweighted) to 42.8% (clinical covariates) to 59.3% (clinical covariates plus embeddings): the protective association is partially attenuated toward, but does not reach, the null. **(F)** Forest plot of odds ratios for dexamethasone versus PONV across the three strategies, confirming the antiemetic effect is preserved after surgical-name embedding adjustment. The summary panel contrasts the reference trial with our replication across the three strategies.

Our analysis in patients with diabetes mellitus similarly reproduced dexamethasone’s non-inferiority, mirroring the original trial’s main subgroup analysis. The subgroup results paralleled the results from the full cohort, where 30 and 90 day SSI maintained noninferiority, while superficial SSI again trended higher (Figure S4A). The adjustment of the odds ratio with increasing covariate strategies is shown in Figure S4B. The higher superficial SSI estimate may reflect subtype-specific residual confounding, differences in outcome ascertainment, or multiplicity across SSI definitions rather than a uniform associated infection effect.

The expected antiemetic association of dexamethasone was preserved even after surgical-name adjustment. Dexamethasone-treated patients had lower raw PONV rates (Figure 4C), and adjusted PONV analyses remained protective across modeling strategies in the full cohort and diabetic subgroup (Figure 4F; Figure S4C). Notably, in the diabetes subgroup, adjustment with clinical covariates alone produced a null dexamethasone-PONV estimate, whereas adding surgical-name embeddings recovered the expected protective association. Thus, the dexamethasone replication aligned with the PADDI trial for the primary outcome of 30-day overall SSI safety in the full cohort and diabetic subgroup while retaining the expected protective association with PONV.

### GAP: Gabapentin, Length of Stay, and Postoperative Pain

The GAP replication recovered the original trial’s null length-of-stay finding across all three surgical subgroups only after full adjustment with clinical covariate plus surgical-name embeddings. Gabapentin exposure varied across abdominal, thoracic, and cardiac surgery (Figure 5A), and postoperative length-of-stay distributions differed substantially by subgroup (Figure 5B). Pain trajectories also varied across the first 48 postoperative hours, reflecting subgroup-specific recovery and analgesic context (Figure 5C).

**Figure 5.**
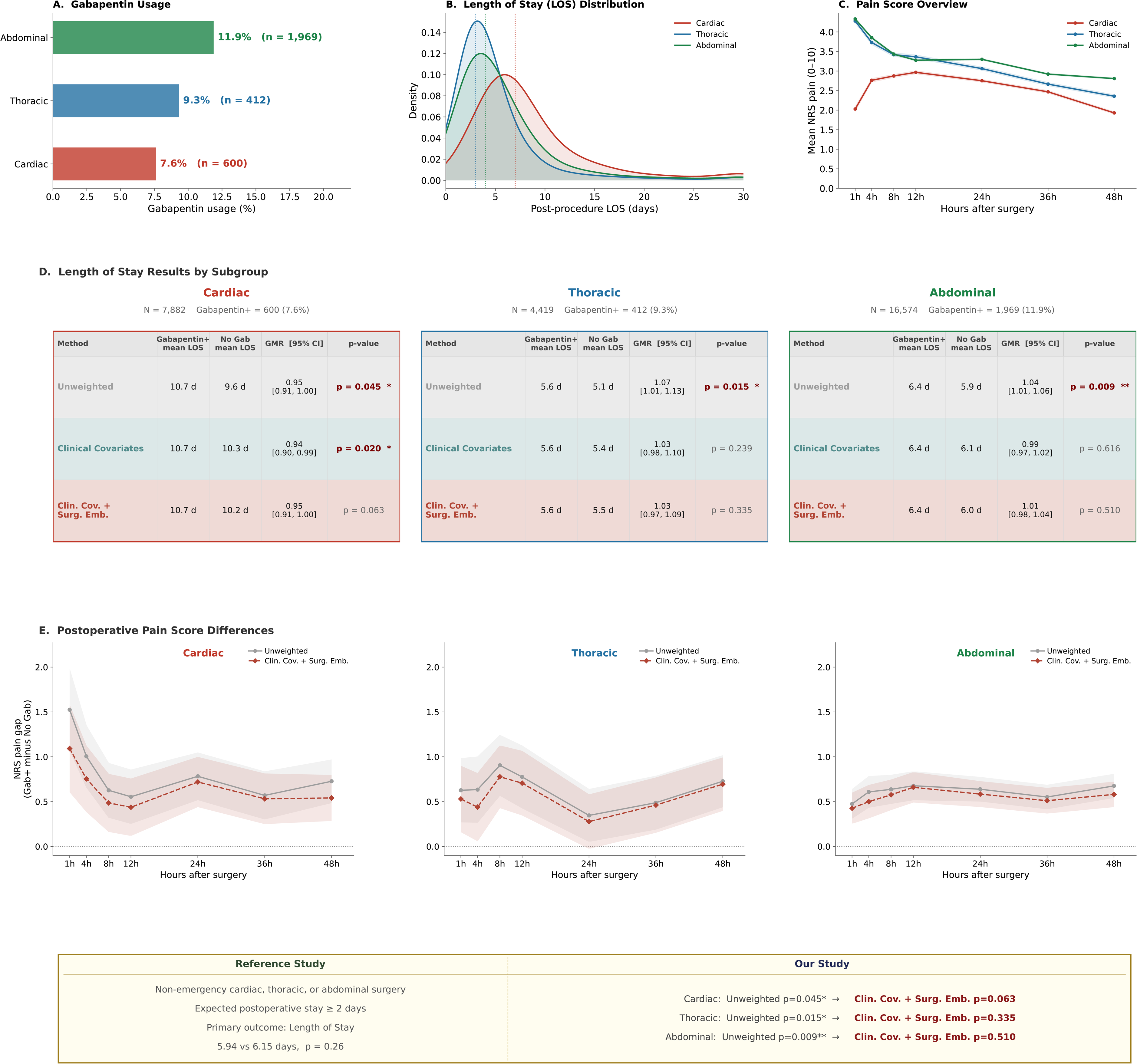
Surgical-name embedding adjustment recovers trial-consistent null length-of-stay findings in gabapentin subgroup analyses. The original GAP study (Baos et al., Anesthesiology 2025) randomized 1,196 patients undergoing major cardiac, thoracic, or abdominal surgery to gabapentin versus placebo; gabapentin did not reduce median length of hospital stay (5.94 vs. 6.15 days; HR 1.07, 95% CI 0.95–1.20, p=0.26). We replicated this analysis across our surgical subpopulations using our three covariate adjustment strategies. **(A)** Gabapentin usage by subpopulation: abdominal 11.9% (n=1,969), thoracic 9.3% (n=412), cardiac 7.6% (n=600). **(B)** Post-procedure length-of-stay distributions by subpopulation (cardiac, red; thoracic, blue; abdominal, green). **(C)** Mean numeric rating scale (NRS) pain scores over 48 postoperative hours by subpopulation. **(D)** Length-of-stay results by subpopulation, reporting gabapentin-exposed and unexposed mean LOS, the geometric mean ratio (GMR) with 95% CI, and p-value under three strategies; the unweighted associations (cardiac 0.95 [0.91-1.00], p=0.045; thoracic 1.07 [1.01-1.13], p=0.015; abdominal 1.04 [1.01-1.06], p=0.009) all show statistical significance that is attenuated to non-significance after clinical covariate plus surgical-name embedding adjustment (p=0.063, 0.335, 0.510, respectively). **(E)** Postoperative NRS pain-score differences (gabapentin minus no gabapentin) across 48 hours for each subpopulation, comparing unweighted (grey) with clinical covariate plus surgical-name embedding (red) estimates. The summary panel contrasts the reference trial’s null length-of-stay finding with the attenuation of each subpopulation’s apparent effect after full adjustment.

In the full eligible GAP-style cohort, gabapentin was not associated with a meaningful reduction in length of stay across adjustment strategies (Figure S5). Unweighted, clinical covariate adjusted, and clinical covariate plus surgical-name adjusted estimates were all consistent with a null effect, aligning with the original GAP trial’s primary finding. However, in subgroup-specific treatment-effect analyses, unweighted and clinically adjusted models suggested length-of-stay associations in some surgical groups, whereas the surgical-name-adjusted estimates were null across abdominal, thoracic, and cardiac surgery (Figure 5D). Postoperative pain-score differences were generally small but remained positive across nearly all time points, indicating higher observed pain scores among gabapentin-exposed patients. Surgical-name adjustment attenuated these differences, consistent with likely residual analgesic selection for patients expected to have greater postoperative pain (Figure 5E). Overall, gabapentin was not associated with a consistent length-of-stay benefit as expected, while demonstrating that surgical-name embeddings were necessary to recover the trial-consistent null result within procedure-specific subgroups.

### Surgical-Name Embeddings Add Additional Value over Control Covariates

The benefit of surgical-name embeddings surpassed the value seen with adding nonspecific covariate dimensions or broad surgical department indicators. Across the three replications, adjustment with department indicators and adjustment with random covariates generally produced estimates that closely resembled clinical covariate adjustment alone (Figure S6). In the GA-CARES replication and cardiac subcohort of the GAP gabapentin study, none of these alternative feature sets recovered the randomized trials’ null results, whereas surgical-name embeddings attenuated both estimates to a nonsignificant association. In the PADDI study, surgical-name embeddings produced a distinct shift in the surgical site infection estimate, while PONV estimates remained stable and protective across feature sets. These patterns suggest that surgical-name embeddings contribute meaningful information not captured by either structured department labels nor the mechanical addition of higher-dimensional covariates.

### Surgical-Name Embedding Sensitivity Analysis

The embedding model sensitivity analysis showed that surgical-name adjustment was not dependent on a single language model representation. We repeated the surgical-name adjustment using BGE-large, MiniLM, and SapBERT embeddings and compared estimates across the three RCT replications for the primary outcomes (Figure S7). Across models, the direction and magnitude of the surgical-name embedding adjusted estimates were broadly similar, supporting that the observed effect of surgical-name adjustment reflected procedure-text information rather than a property of MiniLM alone. These findings support the robustness of surgical-name adjustment across alternative embedding architectures.

The dimensionality-reduction sensitivity analysis showed that conclusions were generally stable across alternative PCA specifications, while also illustrating the importance of retaining sufficient surgical-text information. We repeated the surgical-name embedding adjusted analyses across decreasing numbers of retained principal components to assess the dependency of the results on the primary MiniLM PCA-68 representation (Figure S8). In the GA-CARES analysis, the primary 75% variance specification maintained the trial-consistent null all-cancer TIVA estimate, whereas more aggressive dimensionality reduction to 50% variance produced a spurious protective association, suggesting that excessive loss of surgical procedure information can reintroduce residual confounding. Estimates were otherwise largely preserved across more compact and more expansive embedding summaries down to 75% variance retained, with increasing loss at 50% and more at 25%. This pattern suggests that the primary findings were not driven solely by one PCA cutoff, though adjustment that under-retains embedding dimensions may weaken confounding adjustment in settings where procedure mix strongly influences treatment selection and outcomes.

### Open-Source Software Implementation

We have published a software package at https://pypi.org/project/surgical-embeddings/ to support reuse of the surgical-name embedding workflow. The package accepts free-text operative procedure names as input, generates sentence embeddings, and, if desired, applies PCA matrices trained from our cohort of more than 600,000 surgeries to produce lower-dimensional surgical-text representations (Figure 6). This allows other institutions to leverage the same cohort-derived dimensionality-reduction framework without deriving their own PCA models and supports reproducible adoption of surgical-text adjustment across perioperative datasets.

**Figure 6.**
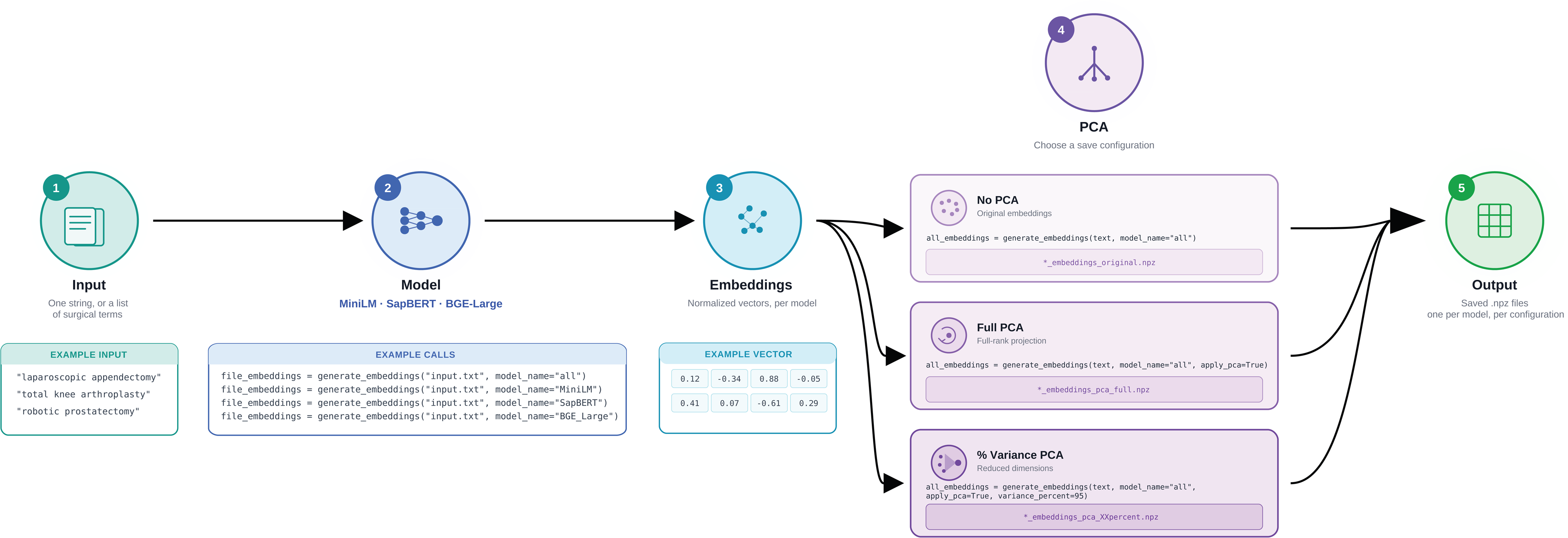
Open-source software workflow for surgical-name embedding generation and dimensionality reduction. The package accepts free-text operative procedure names as input, generates sentence embeddings using the same open-source language models evaluated in this study, and can apply cohort-derived principal component analysis weights trained on more than 600,000 perioperative cases to produce lower-dimensional surgical-text features. This workflow enables other institutions to incorporate surgical-name embedding adjustment into local perioperative observational analyses without deriving their own embedding reduction matrices.

## Discussion

This study presents a surgical-name embedding framework to improve confounding adjustment in perioperative observational research. Using a large perioperative dataset of more than 600,000 surgical cases, we generated dense embeddings from free-text operative procedure names and incorporated them into entropy-balanced observational replications of three recent randomized trials. Across cancer survival, surgical site infection, and postoperative recovery, adding surgical-name embeddings altered treatment-effect estimates in clinically meaningful ways and improved concordance with randomized trial benchmarks. These findings suggest that free-text surgical procedure names encode important procedural context that is not fully captured by standard clinical covariates, and may be beneficial to routinely incorporate into large-scale perioperative observational studies. We also provide software and cohort-derived PCA weights to support external implementation.

We conducted three trial replications to examine multiple settings in which surgical procedure context affected observational inference. In GA-CARES, unweighted and clinical covariate adjusted analyses suggested a protective association between TIVA and two-year survival, producing results discordant with the RCT. This suggested residual confounding, and the original trial’s null result was recovered only after adding surgical-name embeddings. In PADDI, surgical-name adjustment reproduced PADDI’s overall non-harm finding for 30-day surgical site infection while preserving the expected protective association with PONV. Thus, embeddings do not simply attenuate all treatment effects to null, and are able to preserve known, true effects. In the GAP replication, the full cohort aligned with the original RCT’s null result, but spurious significant associations were found in each of the subgroups, which persisted after clinical covariate adjustment in the cardiac surgery subpopulation. The original trial’s null length-of-stay finding was recovered in all subgroups only after surgical-name embeddings were added. Together, these replications suggest that surgical embeddings may have utility for confounding adjustment when treatment selection and outcome risk may be linked to procedure-specific context.

The feature-comparison analysis suggests that surgical-name embeddings added clinically relevant surgical context rather than simply increasing model flexibility. If the main effect of the embedding strategy were due only to higher dimensionality, randomly generated covariates of comparable dimension would be expected to produce similar movement in the estimates. Instead, random covariates, scrambled embeddings, and surgical department indicators largely mirrored clinical-covariate adjustment, and none fully recovered the randomized trial results. The added value of the embedding approach therefore appears to arise from information encoded in free-text surgical names, including distinctions within departments and similarities across departments that are not captured by broad procedural labels.

These findings support using surgical-name embeddings to augment perioperative risk adjustment with routinely available clinical text. Registry-style covariates, including demographic and comorbidity variables, remain important for adjusting for patient-level risk. While tools such as American College of Surgeons NSQIP do incorporate procedure information through structured current procedural terminology codes alongside clinical covariates, coded procedure categories may not fully capture semantic differences between operations, between multiple simultaneous operations, and require a large cohort to have adequate representation of each code. Surgical-name embeddings provide a complementary approach by converting free-text operative names into continuous features that preserve clinically recognizable surgical structure without requiring a manually curated procedure ontology for each analysis.

This study has several limitations. The data came from a single health system, and operative naming conventions, procedure mix, and perioperative practice patterns may differ across institutions. Surgical-name embeddings capture only the information present in the free-text procedure field and may miss important details contained in operative notes, pathology, urgency, surgeon preference, or intraoperative events. The observational cohorts were designed to mirror randomized trials but could not perfectly reproduce trial eligibility, treatment timing, protocol adherence, outcome adjudication, or follow-up. Surgical-name embeddings also cannot address all sources of residual confounding, including unmeasured frailty, contraindications, institutional protocols, or clinician decision-making not recorded in the electronic health record. Sensitivity analyses suggest that confounding adjustment was generally robust to embedding model choice, but the amount of retained PCA information remained important, particularly when aggressive dimensionality reduction removed procedure-context signal.

Overall, this work demonstrates that surgical procedure names contain clinically meaningful information that can improve confounding adjustment in perioperative observational studies. Across three landmark randomized trial benchmarks, surgical-name adjustment helped recover trial-consistent estimates while preserving expected treatment effects. In settings where randomized trials are impractical and procedure-driven treatment selection is common, surgical-name embeddings offer a practical approach for strengthening causal inference from large perioperative datasets.

## Supporting information

Supplemental Digital Content

## Data Availability

A software package has been published and is publicly available at https://pypi.org/project/surgical-embeddings/ and can be used to regenerate intermediate data produced in this work.

## Acknowledgments

Not applicable.

## Clinical trial number and registry URL

Not applicable.

## Prior Presentations

Not applicable.

## Statement

Not applicable.

## Abbreviation Key

BAAI: Beijing Academy of Artificial Intelligence
BERT: Bidirectional Encoder Representations from Transformers
BGE: BAAI General Embedding
CI: confidence interval
GA-CARES: General Anesthetics in Cancer Resection
GAP: Gabapentin for Pain Management after Major Surgery
LLM: large language model
NSQIP: National Surgical Quality Improvement Program
OR: odds ratio
PADDI: Perioperative Administration of Dexamethasone and Infection
PCA: principal components analysis
PONV: postoperative nausea and/or vomiting
RCT: randomized controlled trial
SapBERT: self-alignment pretraining biomedical entity representation model
SSI: surgical site infection
TIVA: total intravenous anesthesia
UMAP: uniform manifold approximation and projection

