## Supplemental Digital Content for "Large Language Model Embeddings of Surgical Procedural Names for Confounding Adjustment in Perioperative Observational Studies"

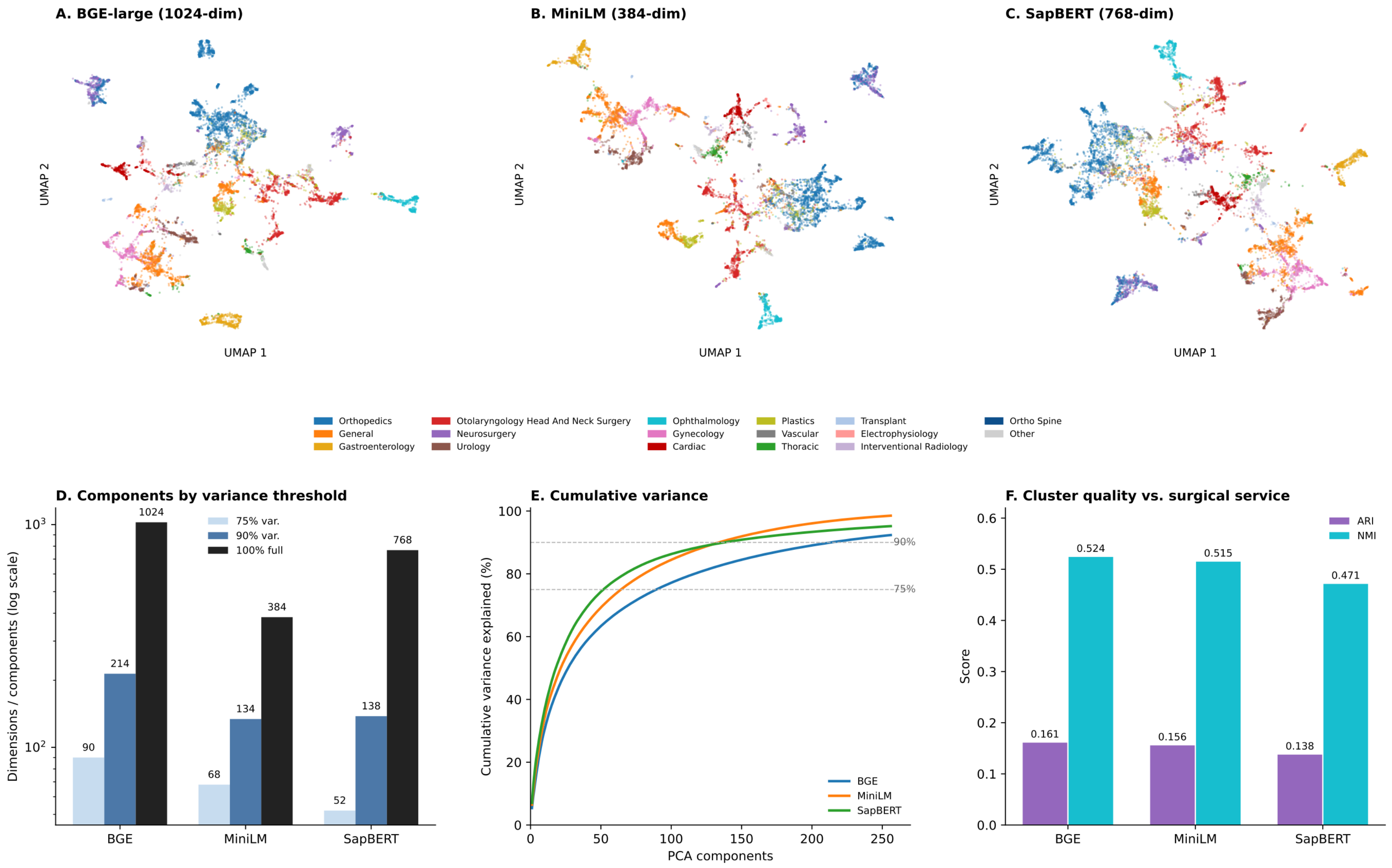


**Figure S1. Embedding-model comparison.** Uniform manifold approximation and projection (UMAP) visualizations of surgical-name embeddings generated with (A) BGE-large, (B) MiniLM, and (C) SapBERT, colored by surgical department. (D) The number of principal components required to retain 75%, 90%, and 100% variance, (E) cumulative variance explained, and (F) clustering agreement with surgical service, assessed using adjusted Rand index (ARI) and normalized mutual information (NMI).

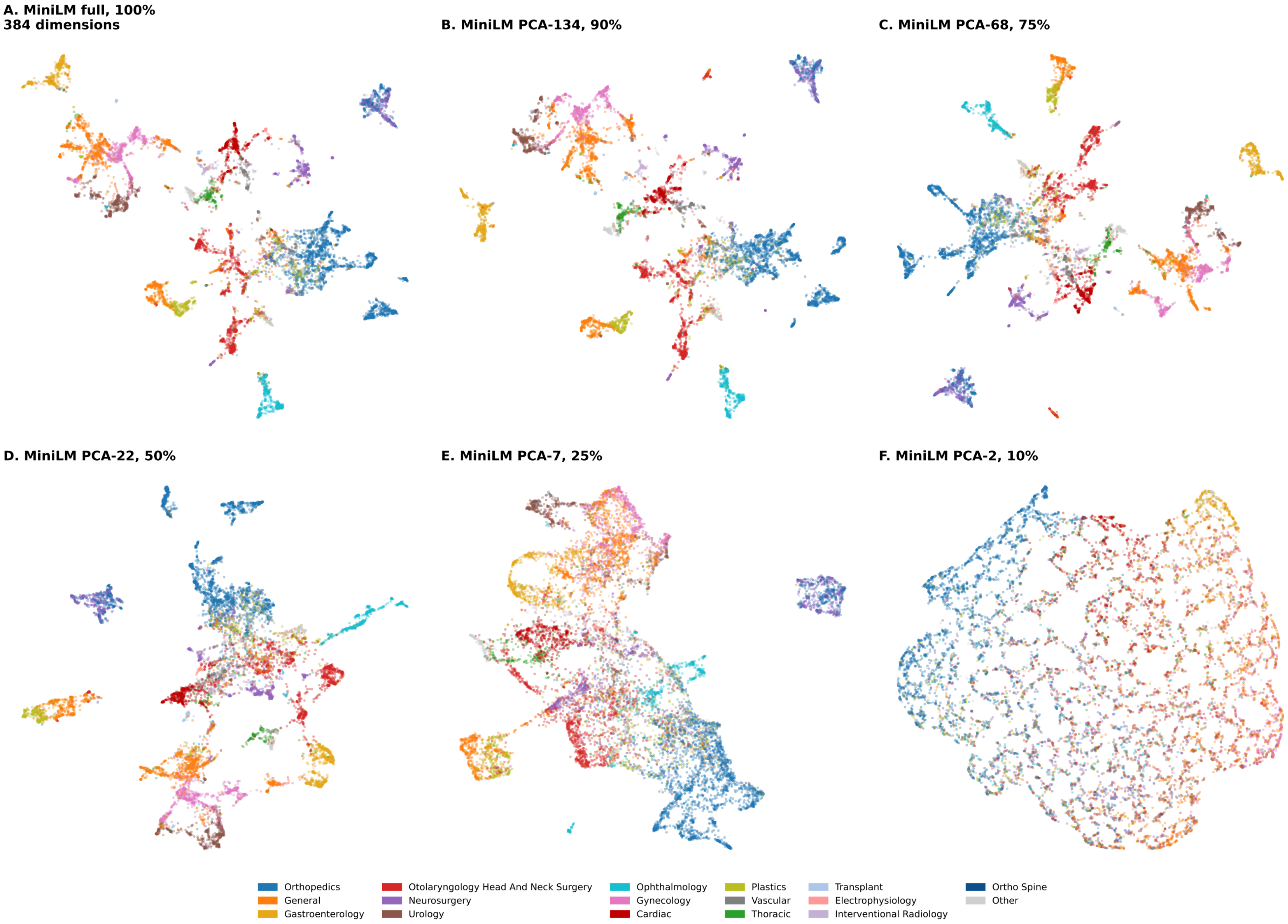


**Figure S2. MiniLM embedding structure across PCA dimensionality thresholds.** Uniform manifold approximation and projection (UMAP) visualizations of MiniLM surgical-name embeddings after progressive dimensionality reduction. Panels compare the full 384-dimensional embedding space with PCA-reduced representations retaining 90%, 75%, 50%, 25%, and 10% of embedding variance.

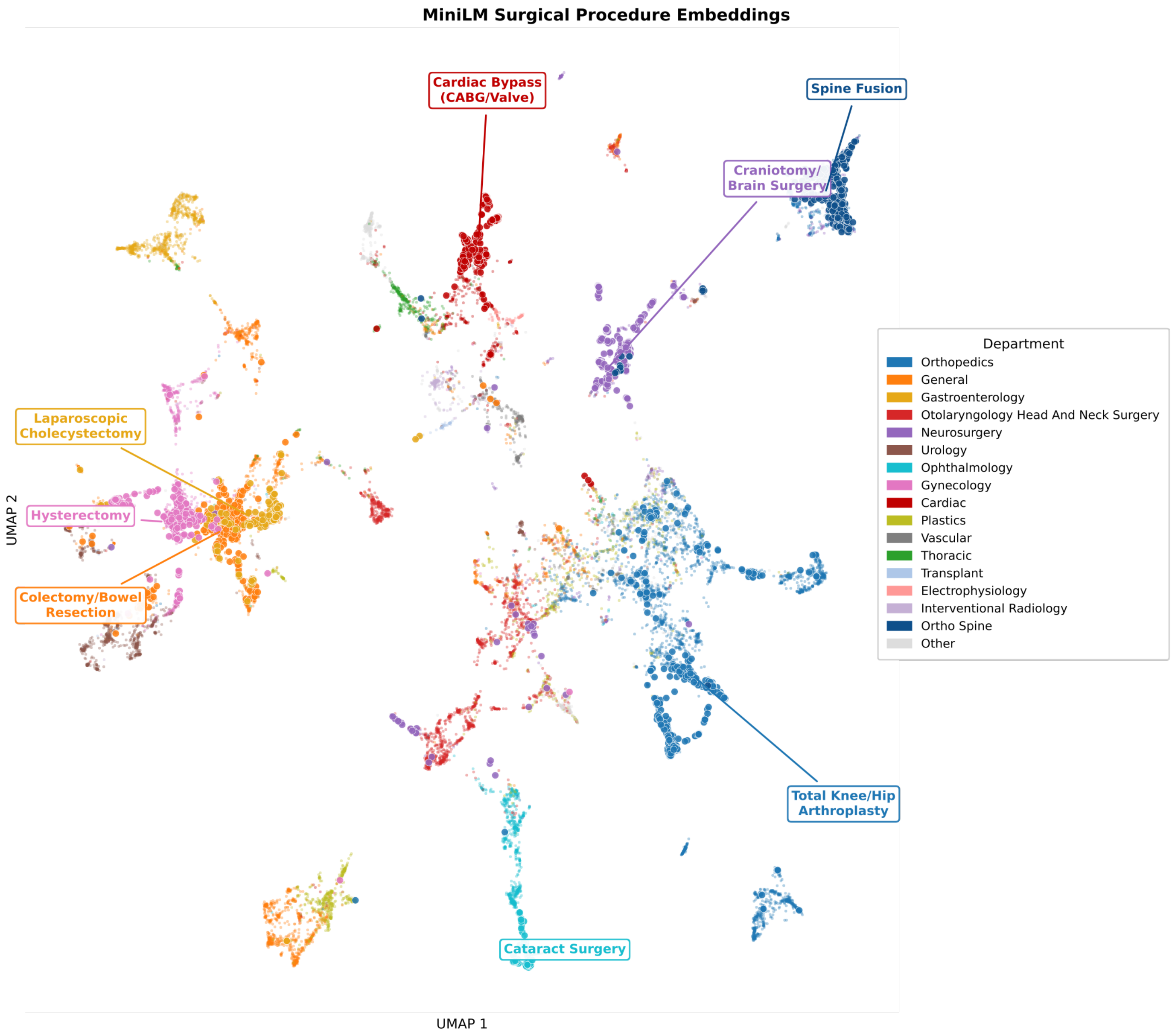


**Figure S3. Labeled MiniLM PCA-68 surgical procedure clusters.** Uniform manifold approximation and projection (UMAP) visualization of the primary MiniLM PCA-68 embedding representation with labeled surgical procedure clusters. The plot illustrates clinically coherent grouping by procedure type, including both department-specific clusters and cross-department procedural similarities.

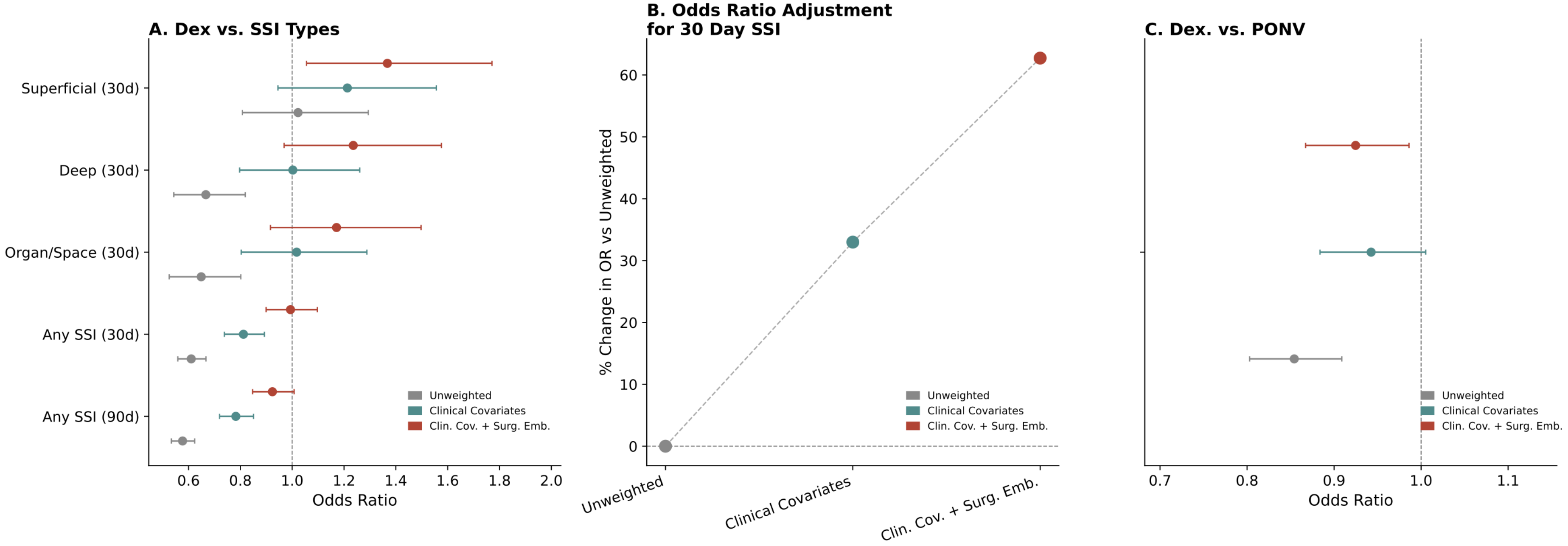


**Figure S4. Dexamethasone analysis in patients with diabetes mellitus.** Subgroup analyses limited to patients with diabetes mellitus, evaluating dexamethasone associations with surgical site infection and postoperative nausea and vomiting. (A) Odds ratios for SSI subtype and time-point estimates, (B) change in the 30-day any-SSI odds ratio across adjustment strategies, and (C) postoperative nausea and vomiting estimates after clinical covariate and surgical-name embedding adjustment.

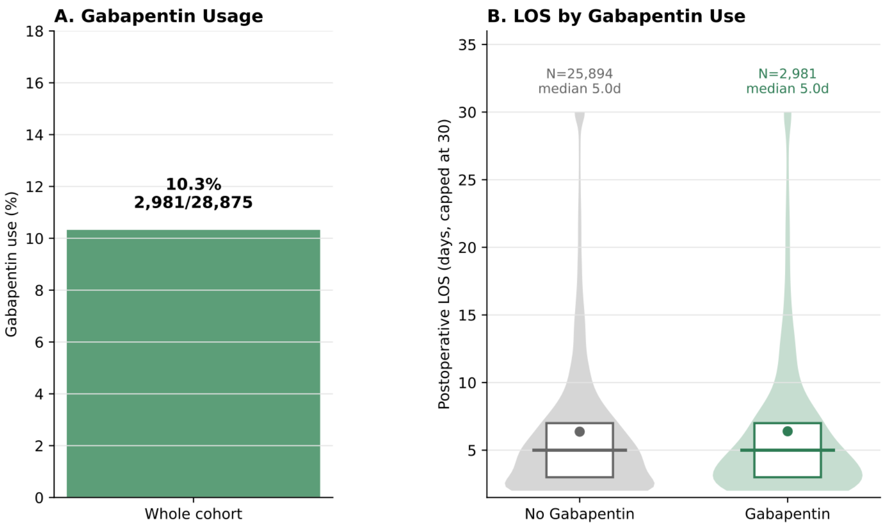


**C. LOS Effect Estimates**

| Method | Gab+ mean LOS | No Gab mean LOS | GMR [95% CI] | p-value |
|---|---|---|---|---|
| Unweighted | 7.1 d | 6.8 d | 0.99 [0.97, 1.02] | p = 0.559 |
| Clinical Covariates | 7.1 d | 6.9 d | 0.99 [0.97, 1.01] | p = 0.467 |
| Clin. Cov. + Surg. Emb. | 7.1 d | 6.7 d | 1.01 [0.99, 1.03] | p = 0.415 |

**Figure S5. Gabapentin full-cohort analysis.** Treatment-effect estimates for perioperative gabapentin in the full eligible GAP-style cohort across unweighted, clinical covariate adjusted, and clinical covariate plus surgical-name embedding adjustment strategies. Length-of-stay estimates were null across unweighted and both adjustment approaches, consistent with the original GAP trial, in contrast to the subgroup-specific analyses where surgical-name adjustment was required to recover trial-concordant null estimates.

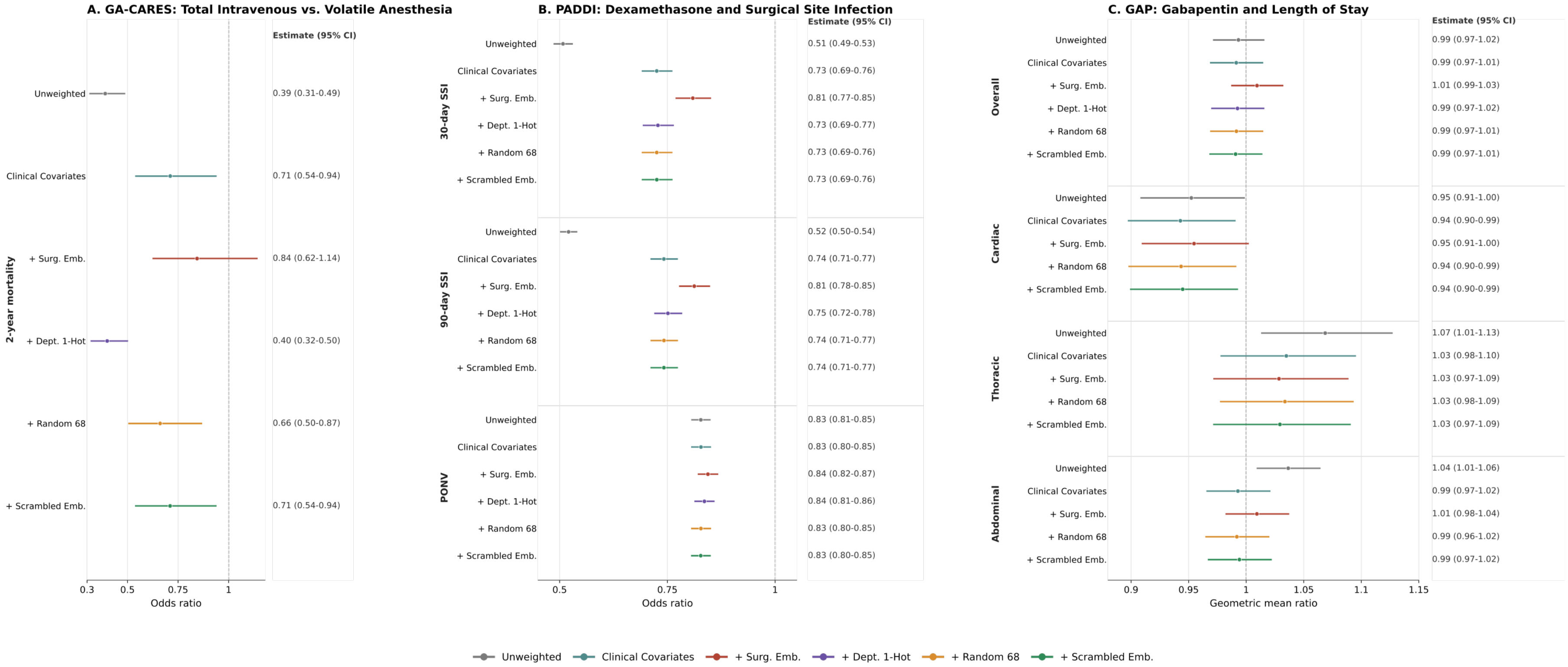


**Figure S6. Feature comparison analysis of surgical context adjustment.** Treatment effect estimates were compared after augmenting clinical covariate adjustment with structured surgical department indicators, randomly generated covariates, randomly scrambled surgical-name embedding components, or the actual surgical-name embedding components. Department indicators, random covariates, and scrambled embeddings largely paralleled clinical-covariate adjustment, whereas observed surgical-name embeddings produced greater estimate shifts and were required to recover all trial results. These findings suggest that surgical-name embeddings contribute procedure-specific information beyond broad service labels or nonspecific high-dimensional adjustment.

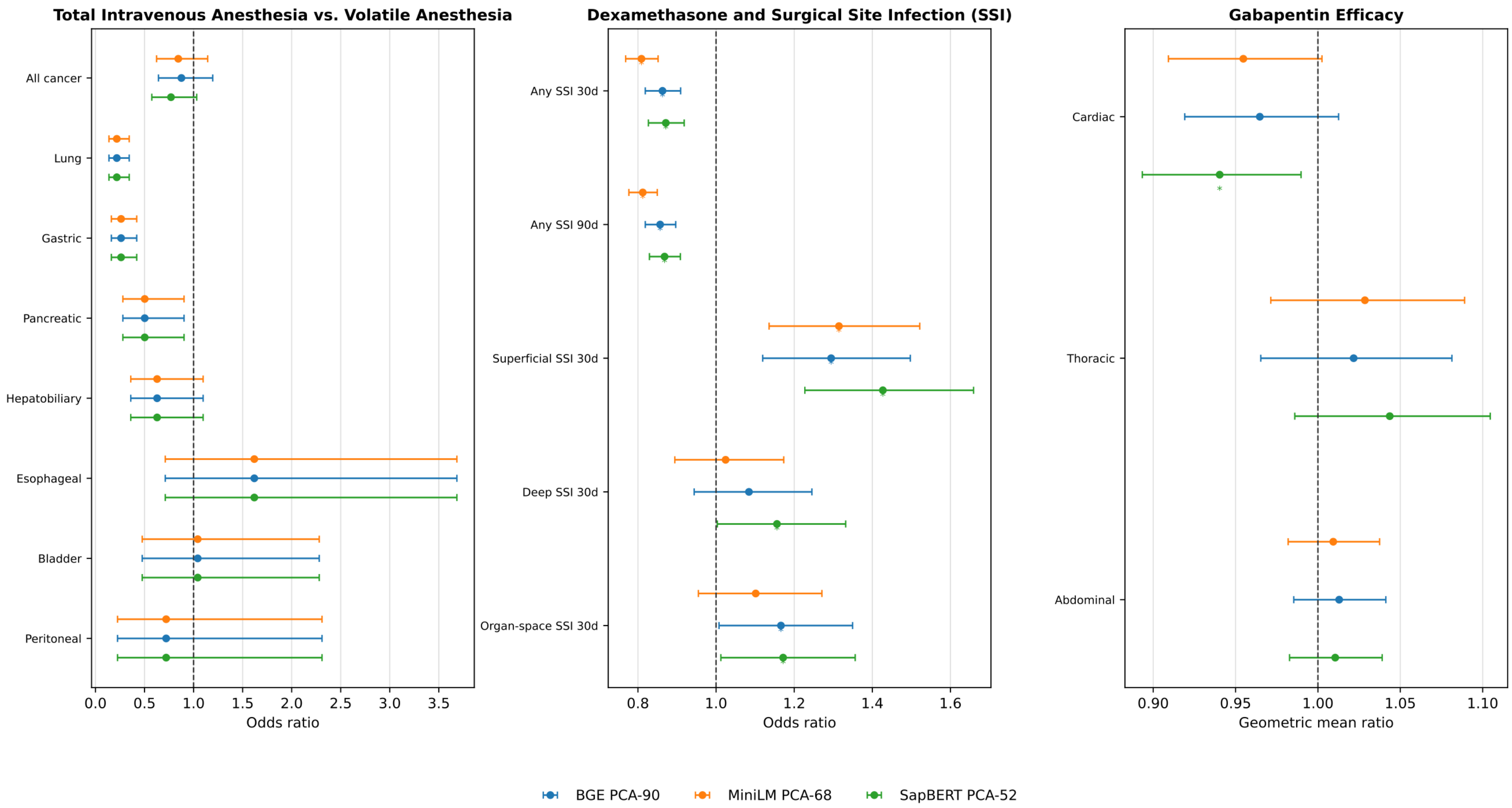


**Figure S7. Embedding-model sensitivity analysis.** Sensitivity analysis comparing surgical-name adjusted estimates generated with BGE-large, MiniLM, and SapBERT embeddings at 75% variance. Panels show treatment-effect estimates for cancer mortality, dexamethasone and surgical site infection, and gabapentin length of stay. Minimal differences are seen across embedding approaches, though only MiniLM maintains the null effect in organ-space SSI at 30 days.

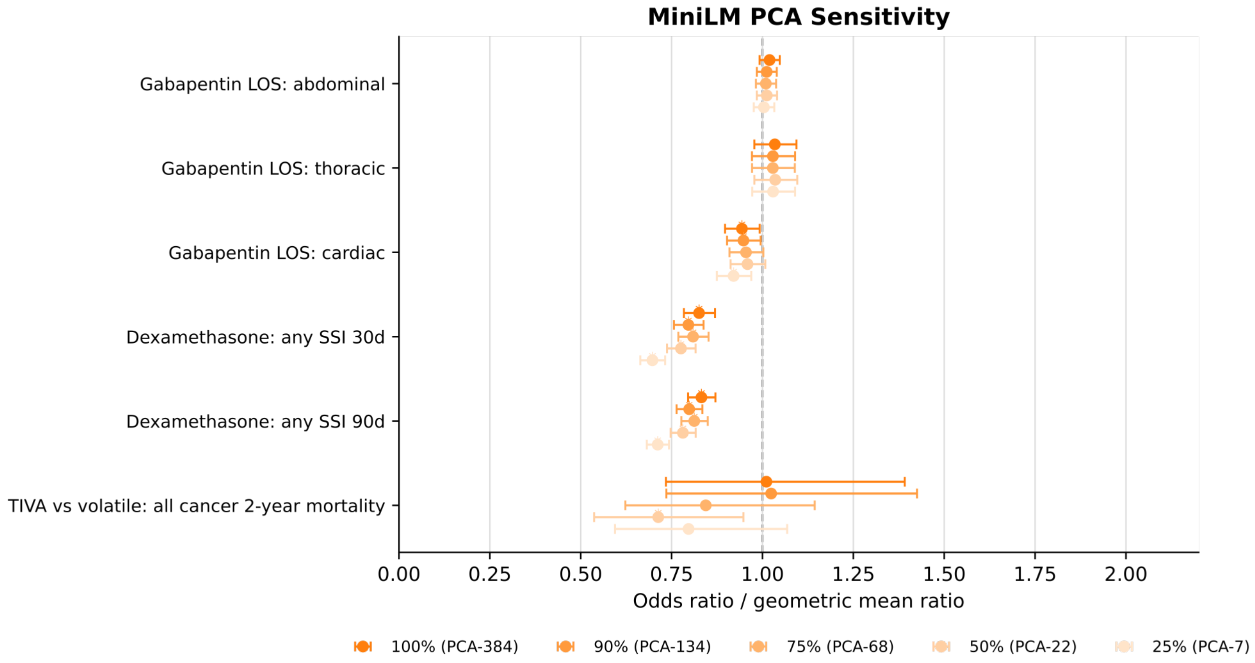


**Figure S8. MiniLM PCA dimensionality sensitivity analysis.** Sensitivity analysis comparing MiniLM embedding-adjusted estimates across alternative PCA variance thresholds. Estimates are shown for the primary outcomes of all-cancer two-year mortality, any SSI at 30 and 90 days, and gabapentin length of stay by surgical subgroup. Loss of adjustment ability is seen with decreasing variance captured, particularly from below 75%.

**Table S1.** GA-CARES Demographics and Outcomes by Anesthetic Exposure

| Covariate | Overall | TIVA | Volatile | p-value |
|---|---|---|---|---|
| Age, years | 57.8 ± 15.7 | 57.5 ± 15.7 | 60.2 ± 15.4 | <0.001 |
| Female sex | 4,299 (58.6%) | 3,935 (60.2%) | 364 (45.6%) | <0.001 |
| Body mass index, kg/m$^2$ | 29.6 ± 8.5 | 29.7 ± 8.6 | 28.6 ± 7.8 | <0.001 |
| ASA physical status | 2.6 ± 0.6 | 2.6 ± 0.6 | 3.0 ± 0.7 | <0.001 |
| Type 2 diabetes | 1,299 (17.7%) | 1,134 (17.3%) | 165 (20.7%) | 0.020 |
| Hypertension | 2,971 (40.5%) | 2,587 (39.6%) | 384 (48.1%) | <0.001 |
| Congestive heart failure | 348 (4.7%) | 264 (4.0%) | 84 (10.5%) | <0.001 |
| Chronic obstructive pulmonary disease | 462 (6.3%) | 378 (5.8%) | 84 (10.5%) | <0.001 |
| Disseminated cancer | 1,034 (14.1%) | 913 (14.0%) | 121 (15.2%) | 0.357 |
| Lung cancer | 2,376 (32.4%) | 2,191 (33.5%) | 185 (23.2%) | <0.001 |
| Gastric cancer | 2,167 (29.5%) | 1,915 (29.3%) | 252 (31.6%) | 0.179 |
| Pancreatic cancer | 816 (11.1%) | 687 (10.5%) | 129 (16.2%) | <0.001 |
| Hepatic cancer | 902 (12.3%) | 798 (12.2%) | 104 (13.0%) | 0.500 |
| Biliary cancer | 264 (3.6%) | 221 (3.4%) | 43 (5.4%) | 0.004 |
| Esophageal cancer | 278 (3.8%) | 205 (3.1%) | 73 (9.1%) | <0.001 |
| Bladder cancer | 593 (8.1%) | 527 (8.1%) | 66 (8.3%) | 0.835 |
| Peritoneal cancer | 214 (2.9%) | 193 (3.0%) | 21 (2.6%) | 0.613 |
| 2-year mortality | 484 (6.6%) | 376 (5.7%) | 108 (13.5%) | <0.001 |

**Table S2.** PADDI Surgical Site Infection Demographics and Outcomes by Dexamethasone Exposure

| Covariate | Overall | Dexamethasone | No dexamethasone | p-value |
|---|---|---|---|---|
| Age, years | 55.6 ± 17.4 | 53.9 ± 17.3 | 59.6 ± 16.8 | <0.001 |
| Female sex | 136,087 (53.6%) | 102,562 (57.6%) | 33,525 (44.1%) | <0.001 |
| Body mass index, $kg/m^2$ | 28.0 ± 6.6 | 27.9 ± 6.5 | 28.2 ± 6.7 | <0.001 |
| ASA physical status | 2.4 ± 0.7 | 2.3 ± 0.7 | 2.7 ± 0.8 | <0.001 |
| Type 2 diabetes | 37,346 (14.7%) | 19,882 (11.2%) | 17,464 (23.0%) | <0.001 |
| Any diabetes | 38,852 (15.3%) | 20,664 (11.6%) | 18,188 (23.9%) | <0.001 |
| Hypertension | 97,862 (38.5%) | 60,330 (33.9%) | 37,532 (49.4%) | <0.001 |
| Congestive heart failure | 18,423 (7.3%) | 8,556 (4.8%) | 9,867 (13.0%) | <0.001 |
| Chronic obstructive pulmonary disease | 15,239 (6.0%) | 8,039 (4.5%) | 7,200 (9.5%) | <0.001 |
| Ascites | 13,138 (5.2%) | 6,180 (3.5%) | 6,958 (9.2%) | <0.001 |
| End-stage renal disease | 6,947 (2.7%) | 1,891 (1.1%) | 5,056 (6.7%) | <0.001 |
| Disseminated cancer | 21,260 (8.4%) | 15,190 (8.5%) | 6,070 (8.0%) | <0.001 |
| Any SSI within 30 days | 8,220 (3.2%) | 4,515 (2.5%) | 3,705 (4.9%) | <0.001 |
| Any SSI within 90 days | 10,842 (4.3%) | 6,040 (3.4%) | 4,802 (6.3%) | <0.001 |
| Superficial SSI within 30 days | 1,126 (0.4%) | 762 (0.4%) | 364 (0.5%) | 0.077 |
| Deep SSI within 30 days | 1,309 (0.5%) | 736 (0.4%) | 573 (0.8%) | <0.001 |
| Organ-space SSI within 30 days | 1,179 (0.5%) | 700 (0.4%) | 479 (0.6%) | <0.001 |
| PONV | 25,544 (10.1%) | 16,961 (9.5%) | 8,583 (11.3%) | <0.001 |

**Table S3.** GAP Gabapentin Study Demographics and Outcomes by Exposure

| Covariate | Overall | Gabapentin | No gabapentin | p-value |
|---|---|---|---|---|
| Age, years | 59.2 ± 15.1 | 58.2 ± 14.9 | 59.3 ± 15.1 | <0.001 |
| Female sex | 13,586 (47.1%) | 1,495 (50.2%) | 12,091 (46.7%) | <0.001 |
| Body mass index, $kg/m^2$ | 28.5 ± 7.1 | 28.7 ± 7.4 | 28.4 ± 7.1 | 0.090 |
| ASA physical status | 2.9 ± 0.7 | 2.9 ± 0.6 | 2.9 ± 0.7 | 0.129 |
| Type 2 diabetes | 5,026 (17.4%) | 723 (24.3%) | 4,303 (16.6%) | <0.001 |
| Hypertension | 13,056 (45.2%) | 1,536 (51.5%) | 11,520 (44.5%) | <0.001 |
| Congestive heart failure | 3,688 (12.8%) | 410 (13.8%) | 3,278 (12.7%) | 0.090 |
| Chronic obstructive pulmonary disease | 1,932 (6.7%) | 302 (10.1%) | 1,630 (6.3%) | <0.001 |
| Disseminated cancer | 3,413 (11.8%) | 552 (18.5%) | 2,861 (11.0%) | <0.001 |
| Preoperative creatinine | 1.0 ± 0.9 | 0.9 ± 0.7 | 1.0 ± 0.9 | <0.001 |
| Cardiac surgery service | 7,882 (27.3%) | 600 (20.1%) | 7,282 (28.1%) | <0.001 |
| Thoracic surgery service | 4,419 (15.3%) | 412 (13.8%) | 4,007 (15.5%) | 0.018 |
| Abdominal surgery service | 16,574 (57.4%) | 1,969 (66.1%) | 14,605 (56.4%) | <0.001 |
| PONV | 2,000 (6.9%) | 247 (8.3%) | 1,753 (6.8%) | 0.002 |
| Acute kidney injury | 3,616 (12.5%) | 396 (13.3%) | 3,220 (12.4%) | 0.185 |
| 30-day mortality | 194 (0.7%) | 22 (0.7%) | 172 (0.7%) | 0.641 |
| Postprocedure Length of Stay, days | 6.9 ± 10.7 | 7.1 ± 14.9 | 6.8 ± 10.1 | 0.255 |
| Pain score at 1 h | 4.0 ± 3.3 | 4.6 ± 3.3 | 3.9 ± 3.2 | <0.001 |
| Pain score at 4 h | 3.5 ± 3.1 | 4.2 ± 3.1 | 3.5 ± 3.0 | <0.001 |
| Mean pain score, 24 h | 3.3 ± 2.0 | 3.9 ± 2.1 | 3.2 ± 2.0 | <0.001 |

**Table S4.** Antibiotic Exposures by Dexamethasone Exposure

| Covariate | Overall | Dexamethasone | No dexamethasone | p-value |
|---|---|---|---|---|
| Cefazolin | 190,700 (75.1%) | 141,017 (79.2%) | 49,683 (65.4%) | <0.001 |
| Vancomycin | 6,217 (2.4%) | 2,816 (1.6%) | 3,401 (4.5%) | <0.001 |
| Cefoxitin | 9,923 (3.9%) | 6,588 (3.7%) | 3,335 (4.4%) | <0.001 |
| Piperacillin/Tazobactam | 8,680 (3.4%) | 3,773 (2.1%) | 4,907 (6.5%) | <0.001 |
| Clindamycin | 10,721 (4.2%) | 7,636 (4.3%) | 3,085 (4.1%) | 0.008 |
| Metronidazole | 12,772 (5.0%) | 10,039 (5.6%) | 2,733 (3.6%) | <0.001 |
| Ceftriaxone | 6,309 (2.5%) | 3,822 (2.1%) | 2,487 (3.3%) | <0.001 |
| Ciprofloxacin | 1,493 (0.6%) | 780 (0.4%) | 713 (0.9%) | <0.001 |
| Gentamicin | 3,047 (1.2%) | 1,608 (0.9%) | 1,439 (1.9%) | <0.001 |
| Ertapenem | 1,658 (0.7%) | 876 (0.5%) | 782 (1.0%) | <0.001 |
| Meropenem | 442 (0.2%) | 97 (0.1%) | 345 (0.5%) | <0.001 |
| Ampicillin | 1,434 (0.6%) | 936 (0.5%) | 498 (0.7%) | <0.001 |
